# Can routine laboratory tests discriminate 2019 novel coronavirus infected pneumonia from other community-acquired pneumonia?

**DOI:** 10.1101/2020.02.25.20024711

**Authors:** Yunbao Pan, Guangming Ye, Xiantao Zeng, Guohong Liu, Xiaojiao Zeng, Xianghu Jiang, Jin Zhao, Liangjun Chen, Shuang Guo, Qiaoling Deng, Xiaoyue Hong, Ying Yang, Yirong Li, Xinghuan Wang

## Abstract

**Background:** The clinical presentation of 2019 Novel Coronavirus (2019-nCov) infected pneumonia (NCIP) resembles that of other etiologies of community-acquired pneumonia (CAP). We aimed to identify clinical laboratory features to distinguish NCIP from CAP.

**Methods:** We compared the ability of the hematological and biochemical features of 84 patients with NCIP at hospital admission and 316 patients with CAP. Parameters independently predictive of NCIP were calculated by multivariate logistic regression. The receiver operating characteristic (ROC) curves were generated and the area under the ROC curve (AUC) was measured to evaluate the discriminative ability.

**Results:** Most hematological and biochemical indexes of patients with NCIP were significantly different from patients with CAP. Nine laboratory parameters were identified to be highly predictive of a diagnosis of NCIP by multivariate analysis. The AUCs demonstrated good discriminatory ability for red cell distribution width (RDW) with an AUC of 0.88 and Hemoglobin (HGB) with an AUC of 0.82. Red blood cell (RBC), albumin (ALB), eosinophil (EO), hematocrit (HCT), alkaline phosphatase (ALP), and white blood cell (WBC) had fair discriminatory ability. Combinations of any two parameters performed better than did the RDW alone.

**Conclusions:** Routine laboratory examinations may be helpful for the diagnosis of NCIP. Application of laboratory tests may help to optimize the use of isolation rooms for patients when they present with unexplained febrile respiratory illnesses.

## Introduction

Chinese people are facing unprecedented panic induced by the outbreak of 2019 Novel Coronavirus (2019-nCov) infected pneumonia (NCIP) in Wuhan, China since December 2019 [1]. Currently, during Spring Festival travel rush, millions of people leave Wuhan city, and the 2019-nCov would spread quickly especially along with people coming out from Wuhan. By 10 Feb 2020, a total of 3,7626 people have been confirmed NCIP in China [2].

The NCIP is considered a relative of the deadly Middle East respiratory syndrome (MERS) and Severe Acute Respiratory Syndrome (SARS) coronaviruses. They are characterized by pneumonia symptoms, such as fever, radiographic evidence of pneumonia, respiratory symptoms and possibly transmitted from animal to human [3-5]. The public health authorities proposed NCIP definitions that combined clinical features (e.g., fever, cough, and anhelation) with epidemiological factors (e.g., travel to a seafood and wet animal wholesale market in Wuhan or direct contact with another patient with NCIP) to improve diagnostic accuracy [6, 7]. Unfortunately, these epidemiological features are not specific and have poor positive predictive value during the outbreak.

The NCIP appears to cause symptoms similar to other etiologies of community-acquired pneumonia (CAP) based on clinical data from 41 NCIP patients [3], and can spread from humans to humans [3, 8]. Distinguishing NCIP from other etiologies of CAP is one of the major challenges of the NCIP outbreak. Despite recommendations that examining hematological and biochemical parameters as part of the diagnostic workup for NCIP [3, 9], it is urgent to evaluate the ability of these features to accurately discriminate cases of NCIP from cases of CAP. Thus, we conducted the current study aiming to evaluate the ability of routine laboratory tests for distinguishing NCIP from other etiologies of CAP and help health workers to effectively, quickly and calmly deal with NCIP.

## Materials and Methods

### Patients and Data collection

To determine the ability of routine laboratory tests, measured at hospital admission, to differentiate NCIP from CAP due to other causes, we compared the hematological and biochemical data of NCIP patients and CAP patients. The 84 NCIP patients presented to our hospital from Dec 26, 2019, to Jan 30, 2020. The patients were laboratory confirmed 2019-nCoV infection by real-time RT-PCR. The CAP group consisted of 316 patients who visited our hospital from January 2018 to December 2018. These patients had ≥2 symptoms and signs of CAP and had evidence of pneumonia revealed by the emergency department physician or internal medicine consultant. Healthy controls included 120 healthy people who made the physical check-up in our hospital from Dec 13, 2019, to Dec 17, 2019. The clinical data collection from patients was approved by the Ethics Committee of Zhongnan Hospital of Wuhan University. Written informed consent was waived by the Ethics Commission for emerging infectious diseases.

### Hematological and serum biochemical examination

Routine hematological and serum biochemical examination was ordered at the discretion of the physicians and were measured using standard methods in our hospital. Fasting whole blood from every patient was collected in an EDTA anticoagulant-treated tube and analyzed within 30 minutes of collection. Routine peripheral blood cells, including hemoglobin, lymphocytes, and monocytes, were analyzed. Routine serum biochemical parameters, including alanine aminotransferase (ALT), aspartate aminotransferase (AST), AST/ALT ratio, total bilirubin (TBIL), direct bilirubin (DBIL), unconjugated bilirubin (UBIL), total protein (TP), ALB, globulin (GLB), γ-glutamyl transpeptidase (GGT), alkaline phosphatase (ALP), and total bile acid (TBA) were measured.

### Statistical analysis

Statistical analyses were conducted using IBM SPSS version 22.0 software. Statistical analysis for the results was performed using the Mann-Whitney U test for only two groups or using one-way analysis of variance when there were more than two groups. The receiver operating characteristic (ROC) curves were generated and the area under the ROC curve (AUC) was measured to evaluate the discriminative ability [10]. Higher AUC were considered to show better discriminatory ability as follows: excellent, AUC of ≥0.90; good, 0.80 ≤ AUC<0.90; fair, 0.70 ≤ AUC< 0.80. A p-value <0.05 was considered statistically significant.

## Results

The mean values of laboratory indexes at the time of hospital admission in NCIP patients and CAP patients were demonstrated in table 1 and figure 1. Both NCIP patients and CAP patients had lower mean lymphocyte counts and platelet counts than healthy control. NCIP patients had significantly lower mean values for WBC, neutrophil, eosinophil, lymphocyte, monocyte, red cell distribution width (RDW), platelet counts, and ALP than did patients with CAP. NCIP patients had significantly higher mean values for hemoglobin, hematocrit, ALB, ALT, and AST than did CAP patients. There were no significant differences in mean values of erythrocyte mean levels of globulin (GLB), γ-glutamyl transpeptidase (GGT), and total bile acid (TBA) between NCIP patients and CAP patients.

**Table 1.** Laboratory values at the time of admission to hospital for NCIP patients and CAP patients.

| Variable | Healthy control<br>(n=120) | CAP<br>(n=316) | NCIP<br>(n=84) | P |
| --- | --- | --- | --- | --- |
| Age (years) | 33(24-39) | 70(55-86) | 58(48-70) | 0.000 |
| Gender (M/F) | 68/52 | 210/106 | 51/33 | 0.148 |
| WBC | 6.46 (5.51-7.45) | 9.1(5.22-10.29) | 5.61(3.59-7.11) | 0.000 |
| NEUT | 3.7(2.88-4.49) | 7.15(3.37-8.59) | 4.25(2.34-5.11) | 0.000 |
| EO | 0.14(0.06-0.18) | 0.15(0-0.15) | 0.02(0-0.01) | 0.000 |
| BASO | 0.04(0.03-0.05) | 0.05(0.01-0.06) | 0.01(0.01-0.02) | 0.000 |
| LYMPH | 2.14(1.79-2.43) | 1.13(0.5-1.48) | 0.91(0.61-1.04) | 0.000 |
| RBC | 4.74(4.32-5.15) | 3.29(2.72-3.84) | 4.1(3.77-4.54) | 0.000 |
| HGB | 145.7(134.2-160.1) | 99.9(83.3-117.1) | 127.6(118.9-138.1) | 0.000 |
| PLT | 237.6(194.25-268) | 191.85(116-253.75) | 159(117-189) | 0.000 |
| MONO# | 0.44(0.34-0.51) | 0.61(0.35-0.77) | 0.41(0.26-0.50) | 0.000 |
| HCT | 42.07(38.95-45.78) | 30.45(25.6-35) | 36.4(33.6-40.7) | 0.000 |
| MCV | 88.81(87.1-92.1) | 92.96(90.13-96.6) | 91.7(89.1-94.3) | 0.000 |
| MCH | 30.76(30.1-32.28) | 30.51(29.5-32) | 31.3(30.2-32.4) | 0.008 |
| MCHC | 345.9(343.1-350.5) | 327.97(323-334.4) | 341(334-346) | 0.000 |
| RDW | 13.46(12.6-13.68) | 15.86(13.8-17) | 13.0(12.2-13.2) | 0.000 |
| MPV | 8.86(8.3-9.4) | 8.87(8-9.5) | 9.9(8.7-10.9) | 0.000 |
| ALB | n.a | 30.09(26.73-33.55) | 35.4(32.3-39.1) | 0.000 |
| TP | n.a | 58.74(52.6-65.52) | 63.5(60.4-67.5) | 0.000 |
| ALT | n.a | 31.67(12-37) | 53.9(18.8-44.3) | 0.000 |
| AST | n.a | 48.77(17.25-39.75) | 64.9(26.8-69.3) | 0.000 |
| GLB | n.a | 28.66(24-32.98) | 28.0(25.3-30.1) | 0.594 |
| GGT | n.a | 60.01(20-66.5) | 52(20-62) | 0.790 |
| TBA | n.a | 8.14(2.43-7.9) | 5.3(2.3-6.4) | 0.557 |
| ALP | n.a | 125.0(73.2-126.7) | 80.0(54.5-84.5) | 0.000 |

NOTE. Data are mean value (interquartile range). Abbreviations: WBC, white blood cell; NEU, neutrophil; EO, eosinophil; BASO, basophile; LYM, lymphocyte; RBC, red blood cell; HGB, hemoglobin; PLT, platelet; MONO, monocyte; HCT, hematocrit; MCV, mean corpuscular volume; MCH, erythrocyte mean corpuscular hemoglobin; MCHC, erythrocyte mean corpuscular hemoglobin concentrate; RDW, red cell distribution width; MPV, mean platelet volume; ALB: albumin; TP: total protein; ALT: alanine aminotransferase; AST: aspartate aminotransferase; GLB: globulin; GGT: γ-glutamyl transpeptidase; TBA: total bile acid; ALP: alkaline phosphatase; TBIL: total bilirubin; DBIL: direct bilirubin; UBIL: unconjugated bilirubin. n.a, not applicable.

**Figure 1.**
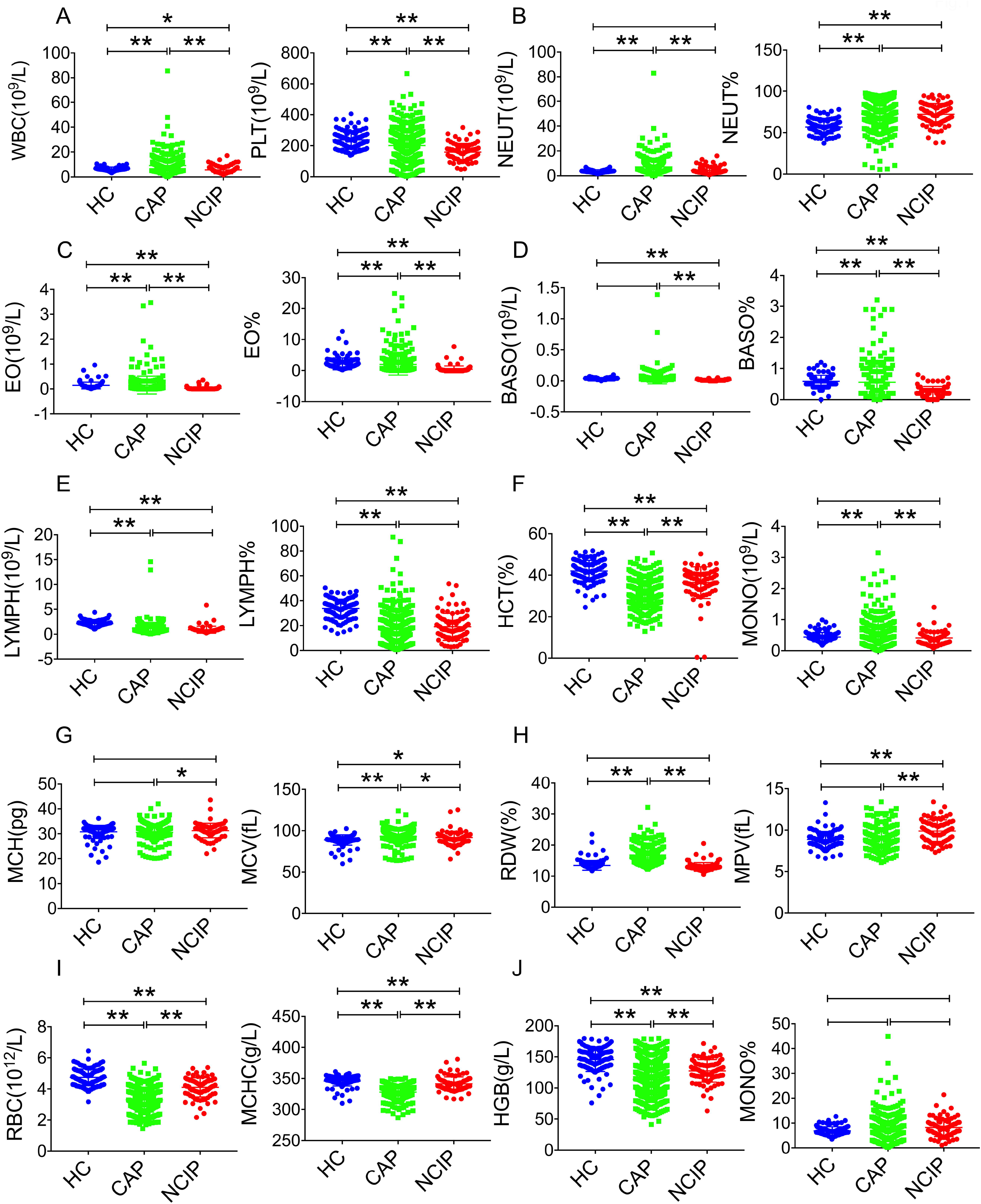
General characteristics of hematological parameters among healthy control (HC), CAP patients and NCIP patients.

The proportion of patients with abnormal laboratory features are presented in table 2. Both NCIP patients (81.0%) and CAP patients (59.5%) had lymphopenia. A significantly higher proportion of NCIP patients presented reduced WBC and eosinophil, normal basophile, and increased AST, whereas a significantly higher proportion of CAP patients had decrease of RBC, hemoglobin, hematocrit, ALB and increase of neutrophil, monocyte and RDW.

**Table 2.**
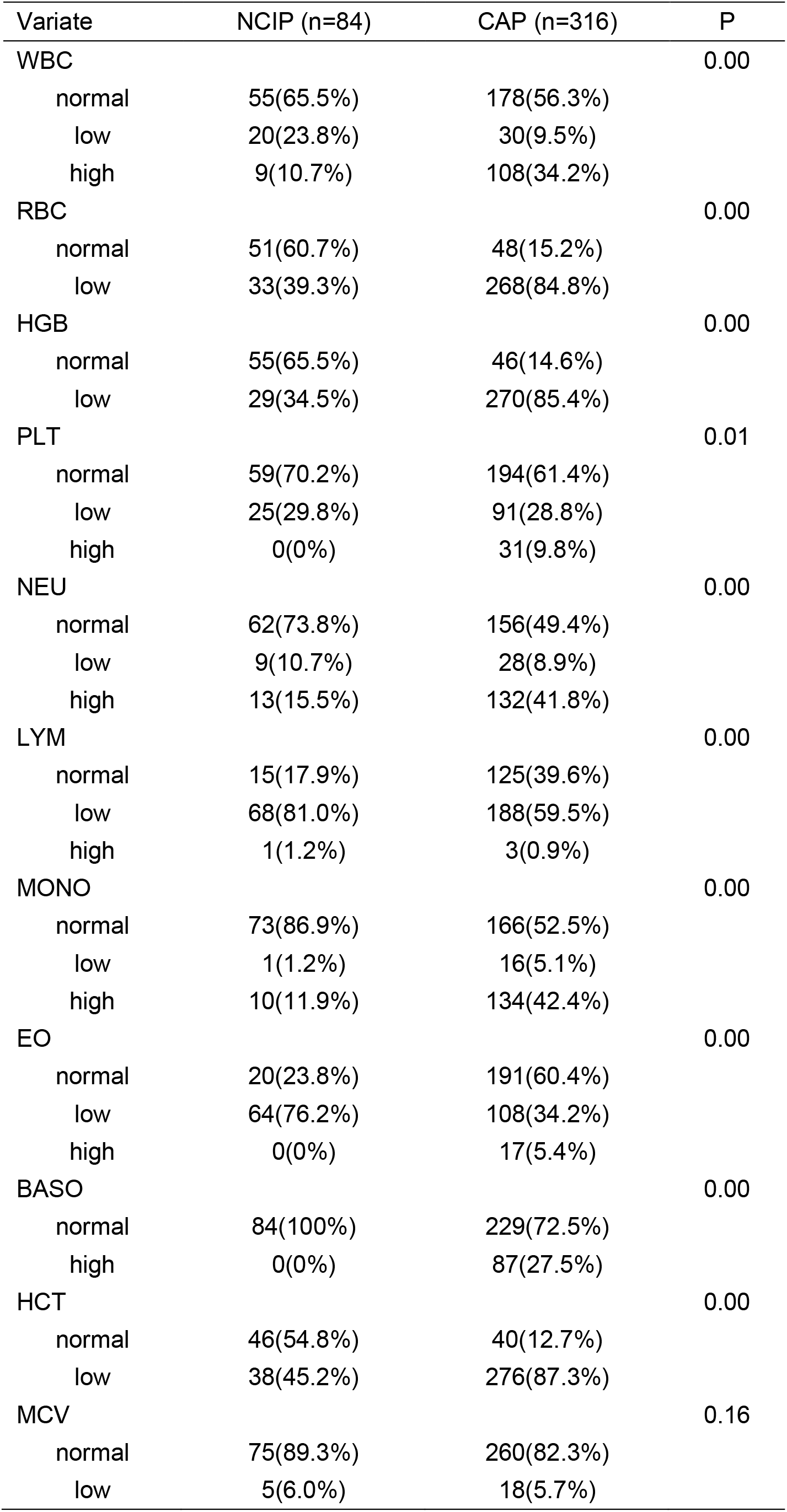
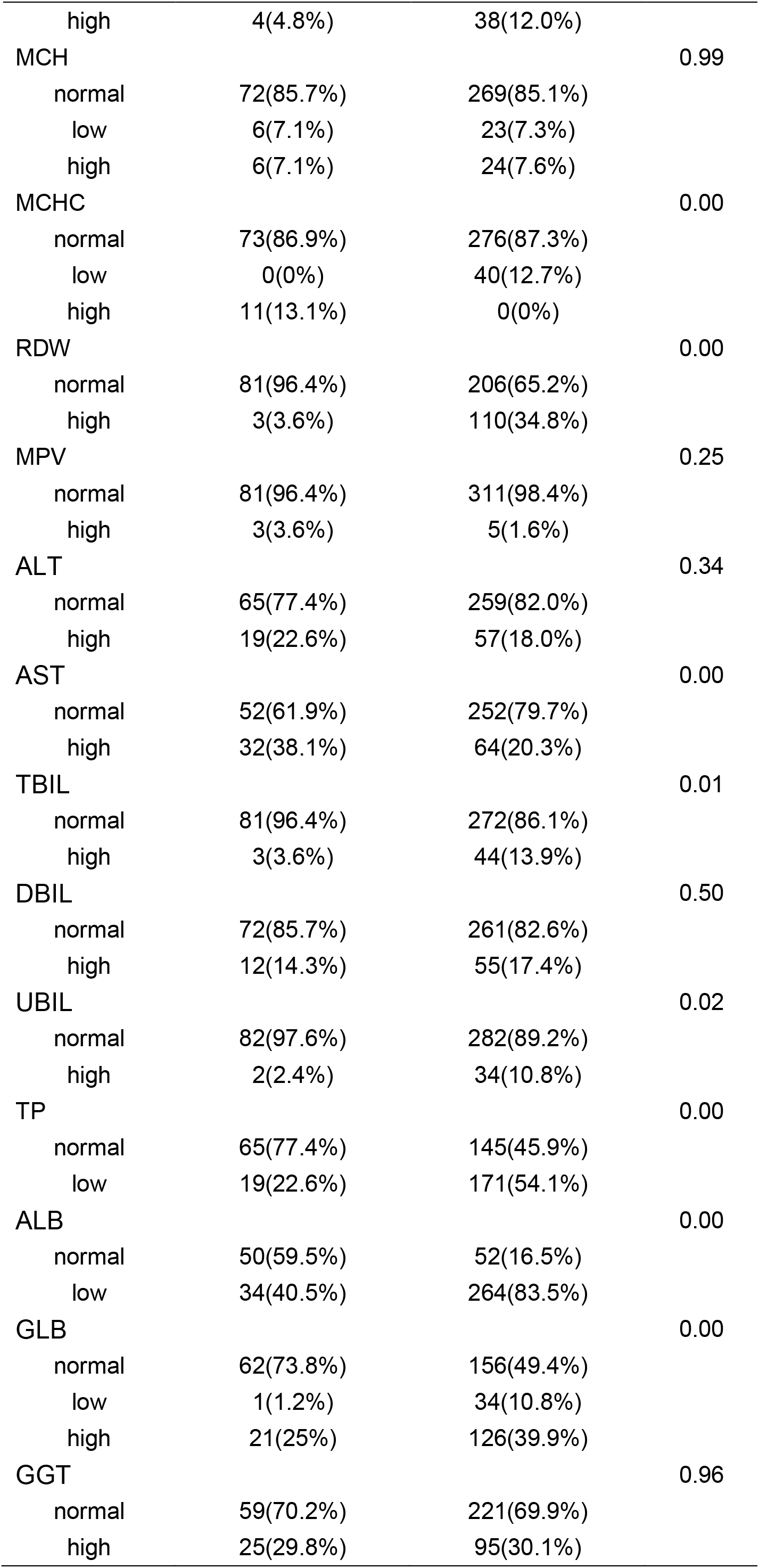
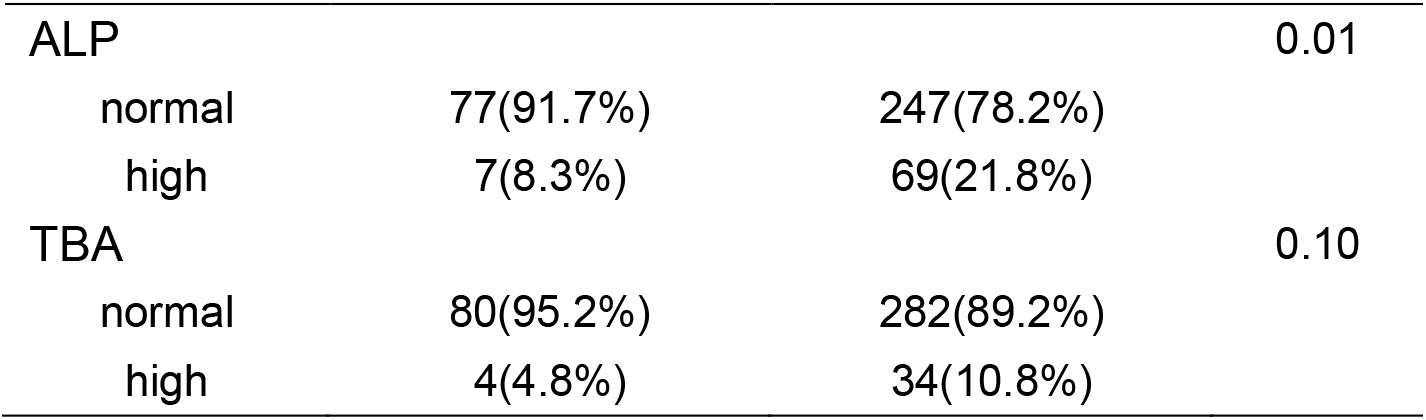
Abnormal laboratory results for NCIP patients and CAP patients.

Multivariate analysis demonstrated the laboratory indexes independently discriminating NCIP from CAP. The OR of the factors to predict NCIP versus CAP were shown in table 3. The ROC curves and AUC (figures 2) demonstrated that RDW (AUC, 0.88) and HGB (AUC, 0.82) had good discriminatory ability. Red blood cell (RBC), albumin (ALB), eosinophil (EO), hematocrit (HCT), alkaline phosphatase (ALP), and white blood cell (WBC) had fair discriminatory ability. Furthermore, the cutoff value for RDW could be 13.35 that provided a reasonable sensitivity (79.8%) or specificity (85.1%). When ROC curves were calculated for combinations of these three parameters, improvement in AUC was presented, with the maximum AUC being 0.90.

**Table 3.** Multivariate predictors of NCIP versus CAP.

| Variate | OR (95%CI) | P |
| --- | --- | --- |
| WBC |  |  |
| normal |  |  |
| low | 0.16(0.03-0.74) | 0.02 |
| high | 4.42(0.98-19.88) | 0.05 |
| RBC |  |  |
| normal |  |  |
| low | 3.41(1.13-10.28) | 0.03 |
| HGB |  |  |
| normal |  |  |
| low | 2.75(0.84-9.01) | 0.09 |
| NEU |  |  |
| normal |  |  |
| low | 26.30(3.71-186.64) | 0.00 |
| high | 2.72(0.71-10.46) | 0.15 |
| LYM |  |  |
| normal |  |  |
| low | 0.25(0.09-0.69) | 0.01 |
| high | 0.07(0.00-3.56) | 0.18 |
| EO |  |  |
| normal |  |  |
| low | 0.21(0.09-0.50) | 0.00 |
| high | 25817931.5131017(0-) | 0.99 |
| RDW |  |  |
| normal |  |  |
| high | 5.84(1.49-22.87) | 0.01 |
| AST |  |  |
| normal |  |  |
| high | 0.29(0.12-0.70) | 0.01 |
| TP |  |  |
| normal |  |  |
| low | 6.22(2.32-16.71) | 0.00 |
| GLB |  | 0.01 |
| normal |  |  |
| low | 9.58(0.79-115.84) | 0.08 |
| high | 4.10(1.54-10.96) | 0.00 |

**Figure 2.**
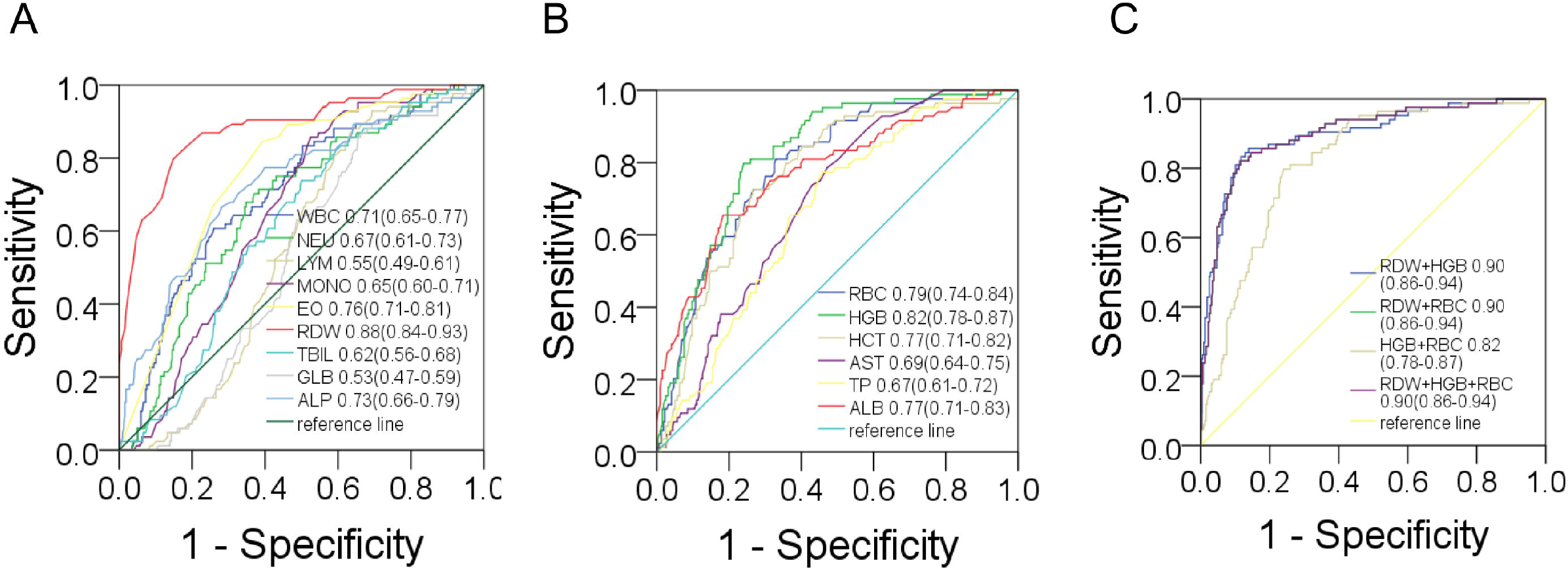
Receiver operator characteristic curves for nine parameters (A), and another six parameters (B), and combinations of the parameters (C), comparing data for CAP patients with that for NCIP patients.

## Discussion

This study indicates that several hematological and biochemical abnormalities occur more frequently in CAP patients than in NCIP patients. The statistically significant difference in mean values was noted for most laboratory features tested except the GLB, GGT and TBA. However, to be useful in diagnosis, the overlap in the distribution of the results for patients with NCIP and those with CAP must be small.

2019-nCov spread quickly in China since the first official announcement in December 2019 by the Wuhan Municipal Health Commission [11, 12]. Possibly, the Spring Festival travel rush that millions of people were on the way home and 2019-nCov would spread fast especially along with people coming out from Wuhan. Because the confirmation diagnosis of NCIP now mainly relies on PCR assays for the detection of 2019-nCov [13], initial medical examination of potential NCIP depends primarily on clinical, radiographic, and epidemiological features [9]. Physicians and public health workers keep struggling with the difficult task of evaluating patients for NCIP presenting with unknown febrile respiratory illnesses. As a result, public health authorities went on revising NCIP definitions to improve the accuracy of diagnosis.

Our findings demonstrated that nine laboratory features independently predictive of discernibility between NCIP and CAP. The identification of the RDW, HGB and RBC count as the best discriminatory ability, with an AUC of 0.89, 0.81 and 0.78, respectively. The discriminatory ability likely results from elevated RBC and HCB while low RDW seen in NCIP. Our study also highlighted laboratory parameters that are common in both NCIP and CAP and therefore not useful in differentiating the 2 diseases. Lymphocytopenia is characteristic and of similar magnitude for both NCIP and CAP. However, lymphocytopenia in NCIP was also accompanied by depletion of EO and normal BASO, whereas it was accompanied by reduced RBC, elevated neutrophil count and monocyte count in CAP.

Because the CAP cohort did not have laboratory indexes over time, trends in laboratory values were unable to perform after hospital admission. Therefore, more significant differences in the laboratory indexes might occur later in the illness, between patients with NCIP and patients with CAP. Our findings indicate that simple laboratory tests may help to distinguish NCIP from CAP. Application of these tests together with epidemiological data may be helpful to avoid misdiagnosis of NCIP as CAP, shorten the time of isolation of patients with respiratory symptoms.

## Data Availability

The datasets generated for this study are available on request to the corresponding author.

## Acknowledgments

We acknowledge all health-care workers involved in the diagnosis and treatment of patients in Wuhan.

## Notes

### Competing Interest Statement

The authors have declared no competing interest.

### Funding Statement

This work was supported by the National Key Research and Development Program of China (2018YFE0204500)

